# TrialScout links published results to trial registrations using a large language model

**DOI:** 10.64898/2026.03.15.26348383

**Authors:** Love von Schreeb, Till Bruckner, Darya Ava Aspromonti, Laura Caquelin, Jamie Cummins, Nicholas J. DeVito, Cathrine Axfors, John P.A. Ioannidis, Gustav Nilsonne

## Abstract

**Background:** Multiple stakeholders need to locate results of registered clinical trials but frequently struggle to find them. Summary results of clinical trials are often not published in trial registries, and publications containing trial results are often not explicitly linked to their respective trial registrations. Finding these results is important to researchers, systematic reviewers, research funders, regulators, clinical practitioners, and patients. **Methods:** We developed TrialScout, a computer program that uses a large language model to match clinical trials registered on ClinicalTrials.gov with corresponding result publications indexed in PubMed. TrialScout’s performance was evaluated through comparison to human-coded matches from previous studies of results reporting rates. Subsequently, TrialScout was applied in a cross-sectional analysis of a random sample of 9,600 completed or terminated trials. **Results:** TrialScout had a sensitivity of 92.5% and a specificity of 81.2% compared to human coders. Manual review of 200 cases where TrialScout disagreed with human researchers showed that a majority (123/200, 61.5%, 95% CI, 54.4–68.3%) of disagreements were due to human errors. When used on 9,600 sampled trials in ClinicalTrials.gov, TrialScout found result publications for 6,110 (63.6%) of trials. **Discussion:** TrialScout reliably located results of completed clinical trials. The tool offers benefits in terms of speed and efficiency. Estimating TrialScout’s accuracy is limited by the lack of a true gold standard. TrialScout can accelerate the process of locating trial results in the scientific literature and can assist in monitoring trial reporting practices.

**Highlights:**

- TrialScout matches published results to clinical trial registrations
- TrialScout uses a large language model to compare publications to registrations
- Accuracy rivals searching by human researchers
- Most disagreements between TrialScout and humans were due to human error
- TrialScout found published results for 63.6% of 9,600 trials from ClinicalTrials.gov

## 1. Background

Clinical trials advance medical research by evaluating the efficacy of interventions. However, advancement of knowledge occurs only if results are reported (1). The Declaration of Helsinki articulates an ethical obligation to publish clinical trial results in a timely manner (2). Non-reporting hinders medical research by biasing evidence reviews (3) and leading to redundant research (1,4), which can expose trial participants to avoidable risks and harms (5) without scientific justification. Bias arises mainly because trials with statistically significant results are more likely to be published, and are published sooner, than trials with null results (6,7). Non-reporting of trial results is thus unethical (8) and can be considered scientific misconduct (3,9).

Despite guidelines and legislation (10,11) promoting timely publication of clinical trial results, results are not always made public (8,12–15). A recent Cochrane systematic review covering 204 research reports on interventional studies registered in ClinicalTrials.gov estimated that only 53% of trials had results published in a journal (7). When completed trials lack results in clinical trial registries, it is often unclear whether results have been published elsewhere. ClinicalTrials.gov sometimes links to publications from trial registrations via researcher submissions and automatic detection of National Clinical Trial numbers (NCT-ID) found in PubMed abstracts and metadata. However, links remain incomplete. In a study of trials conducted by German university medical centres, 50% of registrations did not contain links to their result publications (16). Furthermore, many trial publications do not reference their trial registration numbers (16), despite established guidelines (17–19). Finding trial results often requires searching multiple databases, reading candidate publications, and cross-referencing registration data, which is arduous and time-consuming.

Automated methods to detect results have been developed. Powell-Smith and Goldacre developed TrialsTracker, which detects results published to ClinicalTrials.gov and PubMed publications mentioning the trial’s NCT-ID in the abstract or metadata (20). However, TrialsTracker’s reliance on explicit NCT-ID links limits its sensitivity. Smalheiser and Holt developed a logistic regression model that predicts linkage between trial registrations and PubMed articles using metadata similarity, reporting a sensitivity of 84.6% and a positive predictive value of 90.4% (21). Goodwin and colleagues developed a neural network approach for the same purpose (22). The Smalheiser-Holt model is reported to perform better, but takes several minutes to run per registration since it performs pairwise comparisons across many publications (21). As the PubMed database grows, this computational cost increases, limiting scalability. Appendix 2 gives an overview of currently available tools for automated results detection.

Despite the growing body of evidence of non-reporting of trial results, the extent of the problem is known only for subsets of trials that have been studied, not necessarily representing the population at large. Furthermore, most previous studies have relied on labour-intensive manual searches to find published results, limiting sample sizes. In this study we therefore developed and evaluated a novel tool for the discovery of published clinical trial results. Subsequently, we applied this tool to a large random sample of completed or terminated trials registered in ClinicalTrials.gov, as a proof-of-principle and to provide initial evidence on overall reporting proportions. We also investigated how reporting practices vary by trial characteristics. Industry-funded, earlier-phase trials, and smaller trials have all been found to report results less often (7,23–26). Participant sex has not, to our knowledge, been examined as a correlate of reporting practices. We therefore examined how these four characteristics relate to results reporting in our random sample.

## 2. Methods

### 2.1 Definitions

We defined the following operational terms: *Summary results* are tabular, structured results of clinical trials submitted to the trial registry. *Published results* are results of a clinical trial contained in a peer-reviewed scientific publication, in a thesis, preprint, or published conference abstract. *Reported results* are results of a trial available as summary results and/or published results. This definition of what constitutes results publication is adapted from our previous study on result reporting across the Nordic countries (24).

### 2.2 Development of TrialScout

We developed an automated tool, TrialScout, using Node.js (version 22.7.0) (27). TrialScout retrieves trial registration metadata, searches for candidate publications using a predefined algorithm, and prompts a large language model (LLM) with both the trial registration and publication abstracts to determine whether any candidate publications report trial results. This filters irrelevant candidate publications and distinguishes result publications from other publication types such as study protocols, systematic reviews, and meta-analyses. TrialScout was designed to find *published results* indexed in PubMed for clinical trials registered on ClinicalTrials.gov, EU Clinical Trials Register (EUCTR), and the German Clinical Trials Register (DRKS). These three registries are those covered by the human-curated datasets used for validation. A key constraint of TrialScout is that it can only find results in PubMed with an available abstract.

TrialScout was iteratively designed. Prototypes were run on a subset of the *validation dataset* (described below) constituting <10% of the total dataset. Search strategies and LLM prompts were refined based on manual review of misclassifications. Development concluded when preliminary tests showed high agreement with human data (approximately 85-90% sensitivity and specificity). Figure 1 illustrates TrialScout’s technical implementation. A web version is freely available at https://metaresearch.se/trialscout, and further technical details are provided in Appendix 1 and the source code (https://github.com/lahnstrom/trialscout). TrialScout used OpenAI’s LLM GPT-5.1 (Version “gpt-5.1-2025-11-13”) (28) for all inferences, with temperature set to 1.0 and the reasoning effort parameter set to “medium”. The average cost was 0.043 USD per trial. Runtime varied depending on external API rate limits, hardware, and model settings. TrialScout processed the validation dataset with 5,774 trials in approximately one day on an M2 MacBook Air with 8GB of RAM.

**Figure 1.**
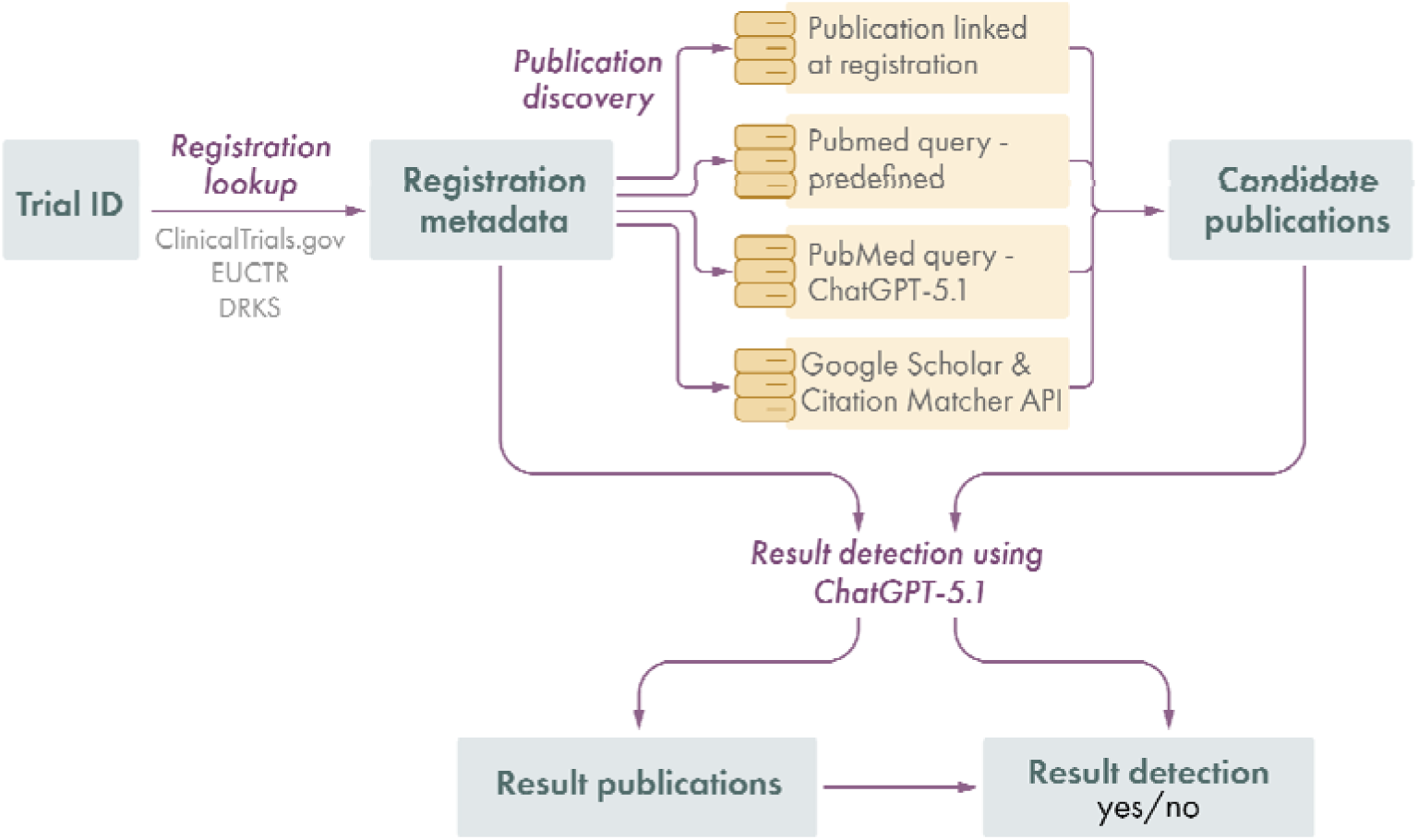
Overview of TrialScout’s result detection algorithm. Dark blue rectangles represent data. Light blue ellipses represent processes. Light grey cylinders represent search strategies. Arrows represent the flow of data. Abbreviations: API: Application Programming Interface; GPT: Generative Pre-trained Transformer; EUCTR: EU Clinical Trials Register; DRKS: German Clinical Trials Register.

### 2.3 Validation of TrialScout Against Human Assessment

We constructed a *validation dataset* with human-curated data as a reference standard, by merging the IntoValue dataset of German clinical trials from 2009–2017 (N=3,790) (25,29) and the dataset from our previous study on Nordic trials completed between 2016–2019 (N=2,112) (24,30). Trials were registered on ClinicalTrials.gov (Nilsonne et al.: N=2,007, IntoValue: N=3,132), EU Clinical Trials Register (EUCTR) (Nilsonne et al.: N=431), and the German Clinical Trials Register (DRKS) (IntoValue: N=658). In the Nordic dataset, 326 trials were cross-registered in both EUCTR and ClinicalTrials.gov; each was classified under the registry of its primary identifier in the source dataset (174 under EUCTR, 152 under ClinicalTrials.gov). After removing 128 duplicates, the validation dataset contained 5,774 registrations.

The IntoValue and Nordic datasets were selected for their data quality rather than their national origin. Validation data had been collected through rigorous processes detailed in the source studies (24,25), where at least two independent researchers conducted manual searches of PubMed and Google (i.e. Google web search) to find trial results, with discrepancies resolved by consensus (24,25). In the dataset from Nilsonne and colleagues, reviewers fully agreed on 75% of trials, while in 11% of cases one reviewer found a publication that the other missed (24). Researchers were instructed to locate only the earliest available result publication, meaning that there is no full publication record for each trial. The validation dataset also contained only pooled consensus judgments rather than per-reviewer data. The definition of a published result varies slightly between the two datasets: the IntoValue dataset included peer-reviewed journal articles and dissertations (25), while the Nilsonne et al. dataset additionally included preprints, congress abstracts, and letters to the editor (24). However, peer-reviewed journal articles make up the vast majority of identified publications in both datasets. In total, 74.4% (n=4,295) of trials had reported results in any format, and 72.5% (n=4,186) had an identified publication.

TrialScout was applied to all 5,774 trials in the validation dataset, and its findings were compared to the human-collected data. Performance characteristics were calculated, including sensitivity, specificity, positive and negative predictive values, and F-score. A true positive was defined as the identification of *any* valid result publication for a trial, rather than the exact one found by humans, since many trials had multiple publications whereas the validation dataset contained only the earliest. Publications identified by TrialScout published after the manual search date cutoffs (November 11, 2020, for the IntoValue dataset; February 15, 2023, for Nilsonne et al.) were excluded to avoid penalizing TrialScout for correctly finding publications that appeared after data collection was completed.

To further validate TrialScout’s accuracy, we manually reviewed a random sample of 200 cases where its assessment disagreed with human reviewers. This sample included 100 false positives (publications found by TrialScout but not humans) and 100 false negatives (publications missed by TrialScout). Based on the search manual by Nilsonne et al. (24), two reviewers independently judged whether the publications matched the trial registrations. Any conflicts were resolved by consensus. Each confirmed TrialScout error was then investigated and categorized.

### 2.4 Performance of TrialScout on ClinicalTrials.gov data

We applied TrialScout to a random sample of trials from ClinicalTrials.gov. A total of 571,121 trial registrations were downloaded as a CSV file from the ClinicalTrials.gov website on February 12, 2026 (Figure 2). Only completed or terminated interventional trials were included, as non-interventional studies generally do not fall under the same legal and ethical requirements as interventional trials. Trials completed before September 30, 2022 were eligible, allowing approximately 3.5 years to report results. Completion was defined by the primary completion date, or the study completion date if the primary completion date was not available. Trials lacking a completion or termination date were excluded. From eligible trial registrations (N=220,167), 9,600 were randomly sampled using the “sample_n” function from the R (version 4.2.2) (31) package “dplyr” (version 1.1.4) (32). The sample size was chosen based on a precision-based sample size calculation with a desired 95% confidence interval of ±1% and an estimated proportion of results reporting of 50%, using the “prec_prop” function from the R package “presize” (version 0.3.7) (33).

**Figure 2.**
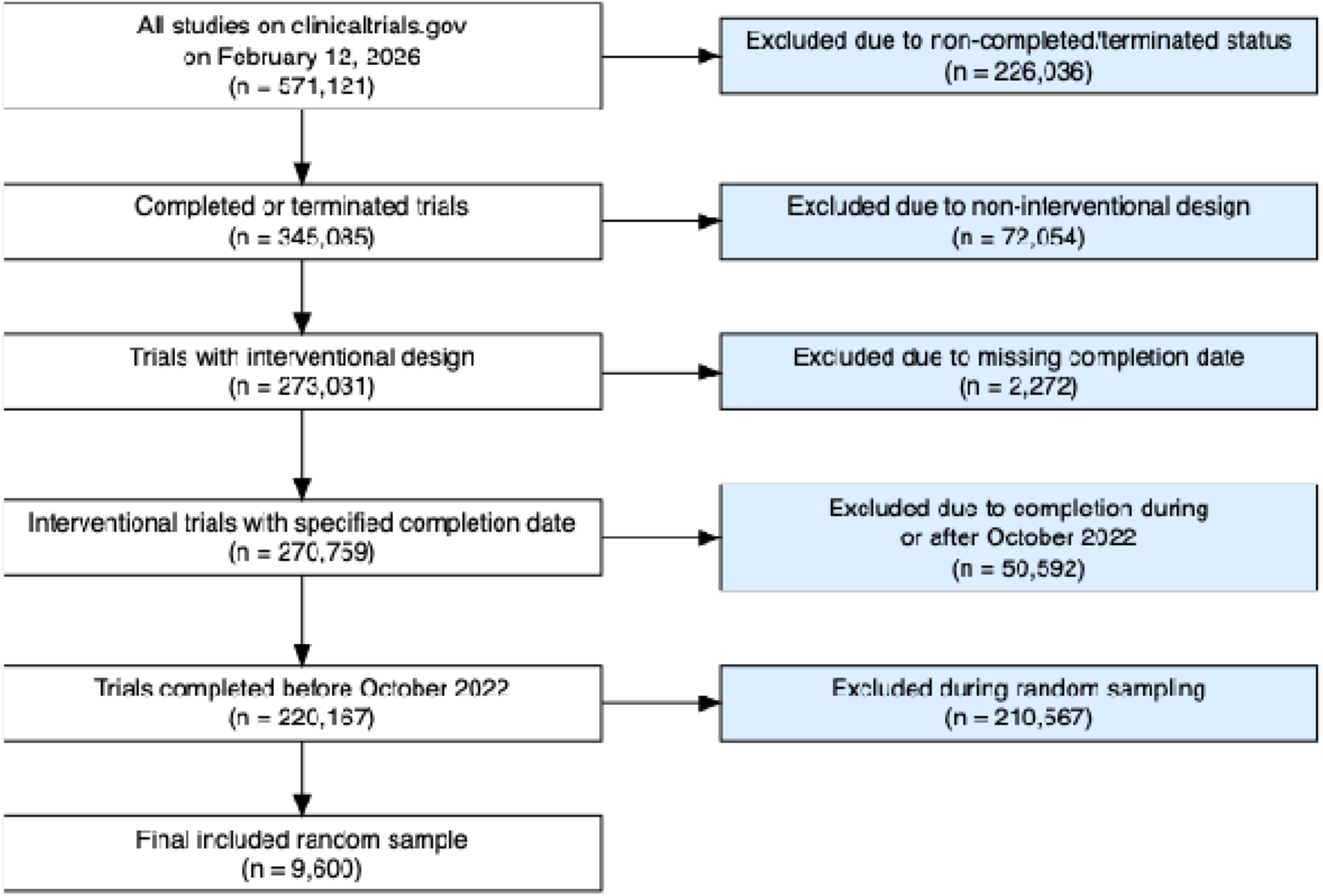
Inclusion flowchart. This figure shows the inclusion and exclusion process for clinical trials retrieved from ClinicalTrials.gov. Only interventional trials with valid date information completed before October 2022 were eligible for random sampling, of which 9,600 were selected.

In February 2026, TrialScout was applied to all clinical trials in the random sample (n=9,600). The primary outcomes were the proportions of *published results*, defined as findings published in a peer-reviewed journal found by TrialScout; and *reported results,* a broader measure indicating that summary results were submitted directly to the trial registry or that published results were found by TrialScout. Data on summary results availability was obtained directly from the ClinicalTrials.gov Application Programming Interface (API). Results reporting proportions for trial subgroups based on phase, completion status, participant sex, and funder type were analysed using descriptive statistics and Pearson’s chi-squared test on published results. Omnibus tests on the full contingency tables were followed by post-hoc chi-squared tests for hypotheses of lower publication proportions among industry-funded, male-only, and early-phase (phase 1 and early phase 1) trials. Separately, univariable logistic regression models examined associations between log-transformed trial enrolment and two outcomes: result reporting and result publication. No covariates were included. R (version 4.2.2) was used for all statistical analyses. The analysis code is available on GitHub (https://github.com/lahnstrom/trialscout) and on Open Science Framework (OSF) (https://osf.io/vh5mr). This cross-sectional analysis is reported in accordance with the STROBE statement (34), and the completed checklist is provided in Appendix 3.

## 3. Results

### 3.1 Validation of TrialScout Against Human Assessment

In the validation dataset, TrialScout performed well against the human reference standard (N=5,774 trials) (Table 1), F-score = 92.7%. The tool detected published results for 72.3% (n=4,173) of trials in the validation dataset, which aligns well with the proportion of trials for which humans found results, 72.5% (n=4,186). Among trials with an associated PubMed-indexed result publication found during manual searches (n=3,914), TrialScout found that publication in 95.4% of cases (n=3,734) and identified that publication as containing results in 91.6% of cases (n=3,585). Performance varied by how the publication was originally found: for trials with results found via links from the registration (n=2,140), TrialScout found results for 98.2% (n=2,101), compared to 86.2% (n=1,668) for trials found through systematic Google search (n=1,934). TrialScout’s agreement with humans varied between registries (Table 1).

**Table 1.** TrialScout validation performance by trial registry.

|  | ClinicalTrials.gov<br>(n=4,837) | EUCTR<br>(n=279) | DRKS (n=658) | All<br>(N=5,774) |
| --- | --- | --- | --- | --- |
| **True positive** | 3,221 (66.6%) | 209 (74.9%) | 444 (67.5%) | 3,874<br>(67.1%) |
| **False positive** | 251 (5.2%) | 9 (3.2%) | 39 (5.9%) | 299 (5.2%) |
| **False negative** | 235 (4.9%) | 13 (4.7%) | 64 (9.7%) | 312 (5.4%) |
| **True negative** | 1,130 (23.4%) | 48 (17.2%) | 111 (16.9%) | 1,289<br>(22.3%) |
| **Sensitivity** | 93.2% | 94.1% | 87.4% | 92.5% |
| **Specificity** | 81.8% | 84.2% | 74% | 81.2% |
| **Positive predictive value** | 92.8% | 95.9% | 91.9% | 92.8% |
| **Negative predictive value** | 82.8% | 78.7% | 63.4% | 80.5% |
| **F-score** | 93.0% | 95.0% | 89.6% | 92.7% |

Performance characteristics of TrialScout when compared to human searches, by trial registry. Abbreviations: EUCTR: EU Clinical Trials Register; DRKS: Deutsches Register Klinischer Studien (German Clinical Trials Register).

To investigate discrepancies between TrialScout and human assessment, a random sample of 200 discrepant cases was manually reviewed by two independent reviewers. Of 100 false positive cases, 79 (79.0%, 95% CI, 69.7–86.5%) were in fact true positives missed by the original manual search (and published before the respective search cutoff dates). For the remaining 21 confirmed false positives, errors were categorized as publications describing a similar but non-matching trial (n=19), or sub-analyses of trial outcomes not listed in the registration (n=2). Similarly, of 100 false negative cases, 44 (44.0%, 95% CI, 34.1–54.3%) were found to be true negatives upon review. For the 56 confirmed false negatives, the primary reason for failure was TrialScout’s search algorithm not finding the correct publication during the search phase (n=51). Of these 51 publications, 51% (n=26) were indexed by PubMed whereas the remaining 49% (n=25) lacked a PubMed entry. The remaining 5 publications were found by TrialScout but classified as not containing results. Overall, across all 200 cases with discrepancies, human error accounted for a larger proportion (123/200, 61.5%, 95% CI, 54.4–68.3%) than TrialScout error.

### 3.2 Random Sample of Completed or Terminated Clinical Trials: Characteristics

TrialScout was applied to 9,600 ClinicalTrials.gov registrations; their characteristics are summarised in Table 2. Enrolment numbers showed a nearly log-normal distribution with a median of 56 participants and an interquartile range of 25-141 participants. The median completion year was 2015. Notably, only 28.6% (n=2,743) of trials had summary results in ClinicalTrials.gov.

**Table 2.** Characteristics of included trials.

| Characteristic | Number of trials (total N=9,600) |
| --- | --- |
| **Study status** |  |
| Completed | 8,561 (89.2%) |
| Terminated | 1,039 (10.8%) |
| **Phase** |  |
| Early Phase 1 | 104 (1.1%) |
| Phase 1 | 1,301 (13.6%) |
| Phase 1/Phase 2 | 331 (3.4%) |
| Phase 2 | 1,505 (15.7%) |
| Phase 2/Phase 3 | 167 (1.7%) |
| Phase 3 | 1,095 (11.4%) |
| Phase 4 | 915 (9.5%) |
| Missing/Not applicable | 4,182 (43.6%) |
| **Completion Year** |  |
| <2005 | 352 (3.7%) |
| 2005–2009 | 1,479 (15.4%) |
| 2010–2014 | 2,573 (26.8%) |
| 2015–2019 | 3,278 (34.1%) |
| 2020–2024 | 1,918 (20.0%) |
| **Enrolled participants** |  |
| 1-99 | 6,294 (65.6%) |
| 100-499 | 2,524 (26.3%) |
| 500+ | 678 (7.1%) |
| Missing | 104 (1.1%) |
| **Summary results** |  |
| No | 6,857 (71.4%) |
| Yes | 2,743 (28.6%) |
| **Participant Sex** |  |
| All | 8,119 (84.6%) |
| Female only | 914 (9.5%) |
| Male only | 561 (5.8%) |
| Missing | 6 (0.1%) |
| **Lead Sponsor Type** |  |
| Other (Universities, Organizations, Networks, Non-U.S. Governmental Agencies, Individuals, Unknown, Ambiguous)<sup>1</sup> | 6,116 (63.7%) |
| Industry | 3,120 (32.5%) |
| U.S. Federal Agency/NIH<sup>2</sup> | 364 (3.8%) |

Characteristics of the 9,600 randomly sampled clinical trial registrations included in the study. Abbreviations: NIH: National Institutes of Health.
<sup>1</sup> Other n=5,823, Non-U.S. Governmental Agencies n=191, Network n=82, Individuals n=17, Unknown n=2, Ambiguous n=1.
<sup>2</sup> NIH n=253, Other U.S. Federal Agency n=111

### 3.3 Identification of Results from Randomly Sampled Trials

TrialScout detected results published in a scientific journal for 63.6% (n=6,110) of a random sample of 9,600 trials. The proportion of trials with reported results, i.e., with publications or summary results, was 72.9% (n=6,998) (Figure 3). TrialScout found 125,528 candidate publications, of which it classified 9% (n=11,256) as results publications. For trials with identified result publications, the mean number per trial was 1.84, with most having a single publication (n=4,163), but some having 10 or more (n=81). Detailed descriptions of each search strategy are available in Appendix 1.

**Figure 3.**
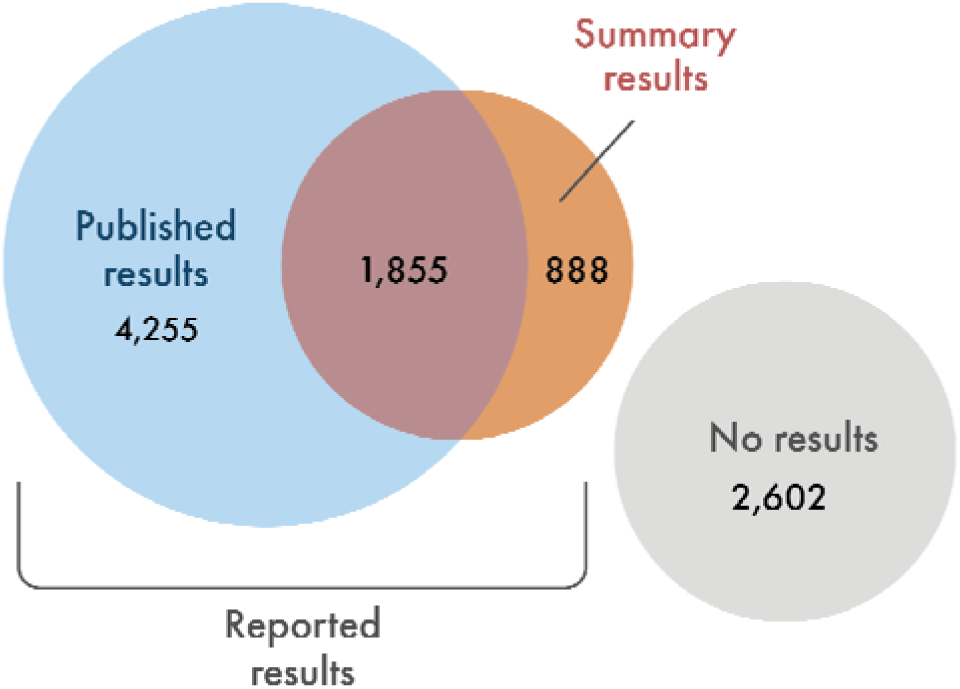
Overlap between published and summary results among 9,600 clinical trials. Published results refer to trial results identified in peer-reviewed publications by TrialScout. Summary results refer to tabular results posted directly on ClinicalTrials.gov. Of the 6,998 trials (72.9%) with any reported results, 4,255 had only published results, 888 had only summary results, and 1,855 had both.

Each doubling of participant enrolment was associated with higher odds of having published (OR=1.31, 95% CI 1.28 to 1.34) and reported (OR=1.22, 95% CI 1.19 to 1.25) results (Figure 4). The relationship between completion year and published results was not monotonic: publication increased from the early 2000s, plateaued around 2010-2015, before declining in recently completed trials, presumably because of more limited time to publish results (Figure 5). Pearson’s chi-squared test showed differences in the proportion with published results across study status, trial phase, and funder type (Table 3, omnibus tests). Post-hoc tests showed that publication proportions were significantly lower for industry-funded trials (55.3% vs. 67.7%, difference -12.4 percentage points, 95% CI -14.5 to -10.3, p<0.001) and for early-phase (phase 1 and early phase 1) versus later-phase trials (53.4% vs. 66.2%, difference -12.8 percentage points, 95% CI -15.8 to -9.7, p<0.001). Trials with exclusively male subjects did not differ significantly in publication proportion (62.4% vs. 63.7%, difference -1.3 percentage points, 95% CI -5.6 to 2.9, p=0.554). Subgroup analyses for all three outcome measures are reported in Appendix 4.

**Figure 4.**
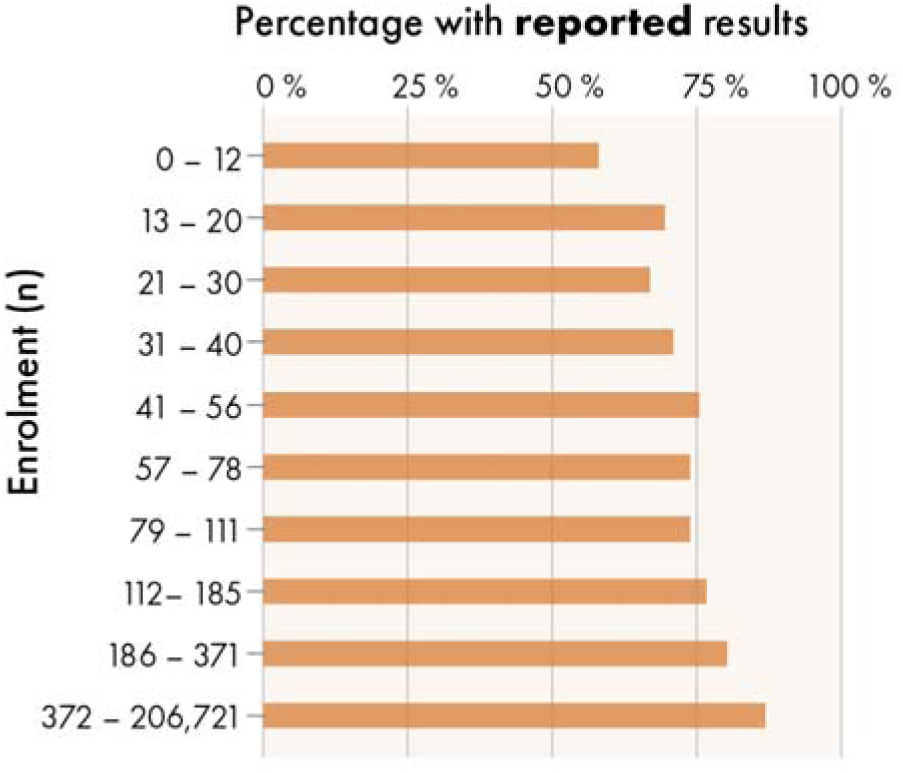
Percentage of clinical trials with reported results by participant enrolment decile. Percentage of clinical trials for which results were reported (either as summary results on ClinicalTrials.gov or in a journal article found by TrialScout) based on the number of participants enrolled in each trial. Sample size is grouped in deciles.

**Figure 5.**
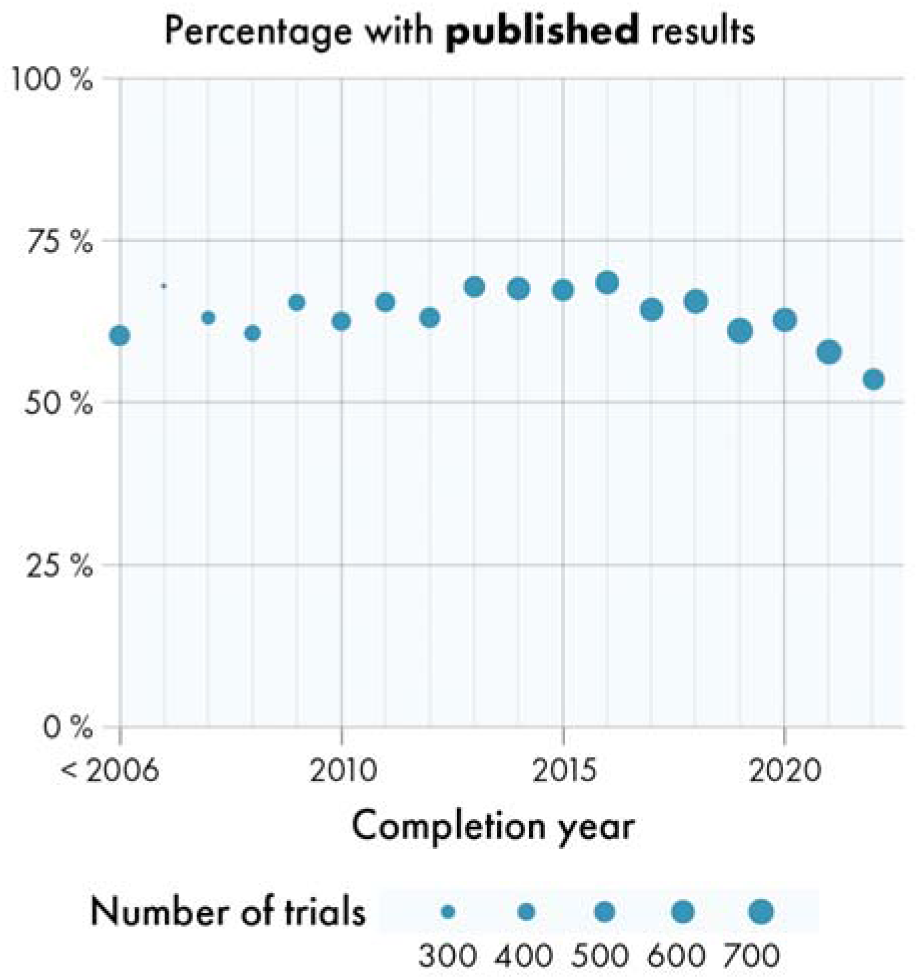
Percentage of clinical trials with published results by completion year. Circles represent years and are sized according to the number of trials completed during that year. Trials from 2005 and earlier have been merged into a single data point as they were comparatively few. Note that while ClinicalTrials.gov launched to the public in February 2000, results reporting has been mandatory only since September 2008.

**Table 3.**
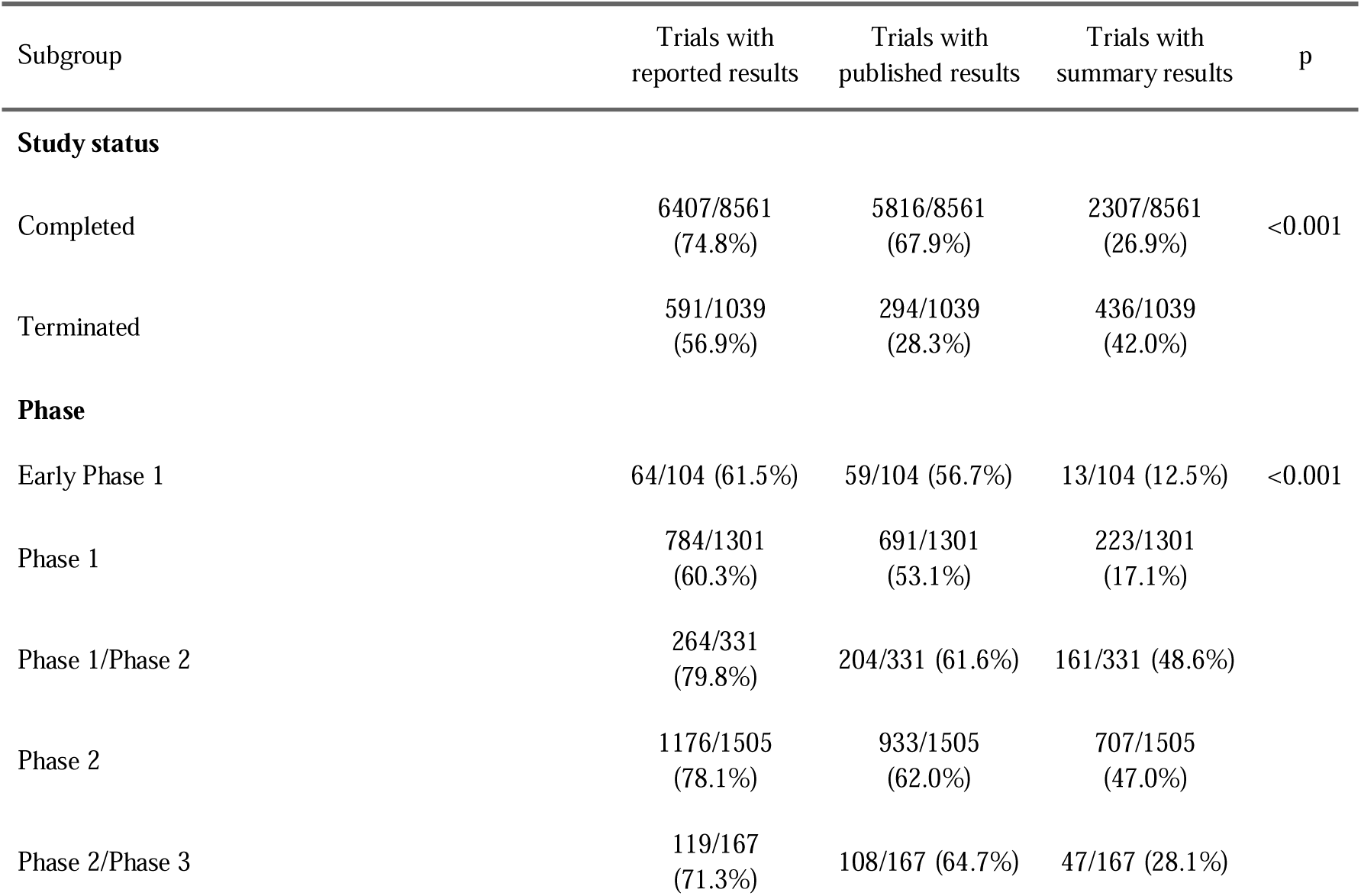
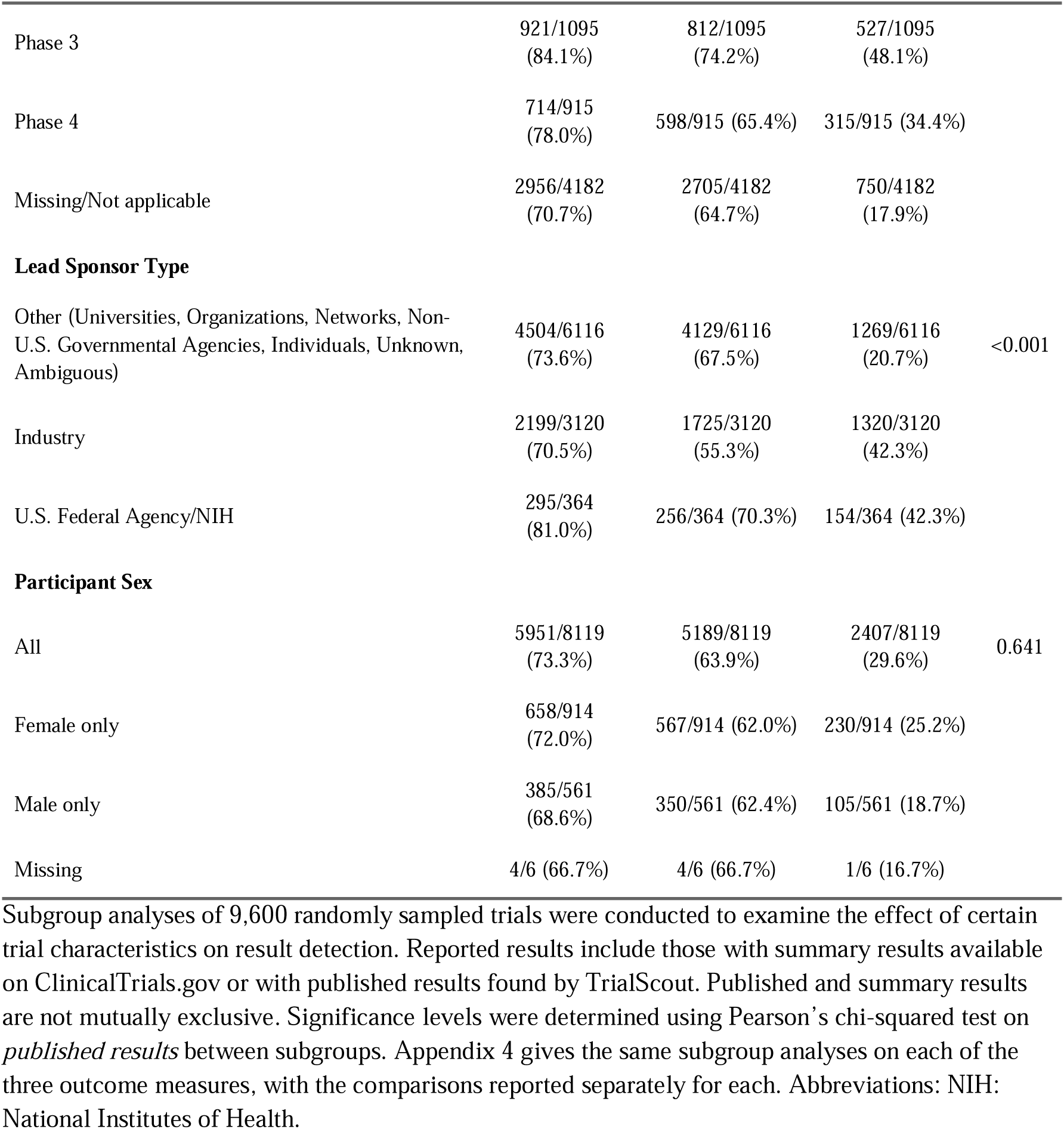
Subgroup analyses for reported, published, and summary results.

## 4. Discussion

TrialScout, our novel tool for linking published results to clinical trial registrations, performs well against a human reference standard. A manual review of disagreement between TrialScout and humans revealed that many of the false positive cases were in fact true positives, and similarly that nearly half of the false negative cases were in fact true negatives. TrialScout’s true performance is therefore likely higher than the performance metrics suggest. When applied to a random sample of 9,600 completed or terminated clinical trials, TrialScout located published results for 63.6% of trials (n=6,110), which can be compared to a recent meta-analysis on clinical trial reporting practices that estimated 53% (7). We found TrialScout to be useful for accurate and scalable linking of clinical trials to published results.

### 4.1 Limitations

A core methodological limitation of this study is our reliance on human researchers’ output when validating TrialScout. We used curated datasets, with data collected through rigorous methods, using multiple independent reviewers (24,25). However, manual review of 200 randomly sampled cases where TrialScout and human reviewers disagreed showed that these datasets contain errors. In four fifths of the cases initially categorized as false positives, TrialScout had in fact found a publication that humans missed. Furthermore, the validation dataset only represents a subset of all trials, not necessarily representative of the entire trial population. The validation dataset is thus best seen as a silver standard for evaluating TrialScout’s performance.

TrialScout has further limitations. Its classifications are not deterministic. Repeating a run can produce different results, and we did not quantify this variability. It searches only PubMed for published results. Results published in journals not indexed by PubMed are thus missed, though searching beyond PubMed generally adds few trials to systematic reviews of interventions (35). Furthermore, publications without abstracts, such as scientific letters, may be missed. While such publications fall out of TrialScout’s intended scope, they must be considered when interpreting our results. Performance was lowest for DRKS, where negative predictive value was 63.4% against 82.8% for ClinicalTrials.gov. Several factors may contribute, including fewer registry-side publication links, dissertations counted as publications in the validation data, and publication in journals that PubMed does not index.

Reporting requirements changed over the period our sample covers. The FDA Amendments Act applied from 2008, its Final Rule and the NIH dissemination policy from January 2017, and phase 1 trials remain largely exempt. Trials completed before and after these dates were thus under different obligations. Furthermore, this study was not prospectively registered, and our results should thus be interpreted with care.

### 4.2 Comparison to previous research

Where existing automated methods rely on NCT-ID searches or metadata matching between trial registrations and publications (20–22), TrialScout uses an LLM to classify publications based on the text of the abstracts. This allows it to use information not present in metadata alone, distinguishing true result publications from protocols, secondary analyses, and reviews. Although an LLM is more computationally intensive per registration-publication comparison, TrialScout achieves faster runtime than the Smalheiser-Holt model (seconds vs. minutes per trial) (21) by combining filtering of the PubMed corpus (its search strategies) with the cloud infrastructure accessible through OpenAI’s API. Due to the different way these previous models have been evaluated, data do not permit a head-to-head comparison to TrialScout.

TrialScout’s high performance allowed us to analyse a large sample of randomly selected clinical trials, making this one of the largest studies on result reporting (7). TrialScout’s analysis gives an overall publication proportion of 63.6%, which is higher than the figure of 53% in Showell and colleagues’ meta-analysis. This discrepancy may reflect differences in the studied trial cohorts, methodologies, or limitations of TrialScout’s accuracy. Furthermore, our sample is limited to trials registered on ClinicalTrials.gov, which likely disproportionately represents trials from the United States rather than the global trials landscape. The higher proportion that we observed may also reflect the fact that older trials (when reported results were less frequent) were more prominently represented in Showell and colleagues (7).

Our subgroup analyses mostly confirmed established findings, with higher publication proportion for trials with high enrolment and higher phases, and lower proportions for industry-sponsored and terminated trials (7,23–26). Trials with exclusively male subjects did not differ in publication proportion. However, our subgroup analyses were univariable and did not adjust for potential confounders such as study size. Additionally, we saw no clear increase in reporting proportions since 2010. We did note a relative decline in the most recent years, likely because many recently completed trials have yet to report their results.

### 4.3 Implications

Our analysis of 9,600 trial registrations demonstrates that TrialScout can be applied at scale, providing a starting point for investigations of reporting practices with humans in the loop. Reporting practices are currently assessed in separate cross-sectional studies of the reporting landscape. Automated linking allows studies to instead be monitored continuously, and at a registry-wide level. Given its high positive predictive value, TrialScout is perhaps best used as a first-pass screening tool, allowing researchers to focus manual searches on trials where TrialScout failed to find results. Furthermore, TrialScout can complement existing monitoring tools such as the FDAAA TrialsTracker (36) or ideally be embedded in the trial registries themselves. We intend to support additional registries. Each requires its own interface, since registries differ in both the data they make available and their structure.

## 5. Conclusion

We show that TrialScout, a novel LLM-based tool, can be used to link published results to clinical trial registrations. The use of AI tools to link research objects may enable a scaling up of efforts to match clinical trial results to registrations, with potential value for metascientists as well as for all stakeholders engaged in clinical trials transparency and evidence synthesis.

## Declarations

### Ethics approval and consent to participate

Not applicable.

### Consent for publication

Not applicable.

### Data availability

The datasets generated and analysed are available in our GitHub repository (https://github.com/lahnstrom/trialscout) and on Open Science Framework (https://osf.io/vh5mr). TrialScout can be accessed at https://metaresearch.se/trialscout.

### CRediT authorship contribution statement

**Love von Schreeb**: Conceptualization, Methodology, Software, Validation, Formal Analysis, Investigation, Data Curation, Writing - Original Draft, Visualization, Project Administration. **Till Bruckner**: Writing - Review & Editing. **Darya Ava Aspromonti**: Validation, Writing - Review & Editing. **Laura Caquelin**: Validation, Writing - Review & Editing. **Jamie Cummins**: Writing - Review & Editing. **Nicholas J. DeVito**: Writing - Review & Editing. **Cathrine Axfors**: Writing - Review & Editing. **John P.A. Ioannidis**: Supervision, Writing - Review & Editing. **Gustav Nilsonne**: Conceptualization, Methodology, Supervision, Writing - Review & Editing.

## Acknowledgements

We thank Meike Latz for expert assistance with preparing figures 1 and 3–5.

This paper originates from Love von Schreeb’s Master Thesis in the medical program at Karolinska Institutet.

## Declaration of generative AI and AI-assisted technologies in the manuscript preparation process

The large language model “Claude” and its associated tool “Claude Code”, provided by Anthropic, were used to assist with development of TrialScout. This included the generation of code snippets, scripts, and documentation. All such generated content was reviewed, tested, and verified by the authors.

