## Appendix 1-2 for "TrialScout links published results to trial registrations using a large language model"

### Appendix 1 – TrialScout’s Technical Implementation

The problem of result detection can be described as twofold; first, from a given trial registration with its associated metadata, publications that are potential matches must be identified from the corpus of all known publications; second, the identified publications must be screened for results matching the trial registration.

#### Data preparation and processing

To facilitate lookup of publications and registrations, data sources were retrieved and stored offline. All registration data available at ClinicalTrials.gov were downloaded using the ClinicalTrials.gov download functionality. The individual trial registrations were saved in a structured text format known as JavaScript Object Notation (JSON).

#### Publication discovery

As of March 2026, there are more than 39 million entries in the PubMed database. Each trial registration may be linked to a subset of these PubMed entries, of which a subset may contain the trial results. TrialScout relies on a computationally expensive machine learning model - an LLM - to predict whether a PubMed entry matches a trial registration. Due to the size of PubMed, applying the LLM to each possible pair of trial registration and publication (i.e., 39 million pairs per trial) would be infeasible. Thus, through an exploratory process, a method of “publication discovery” was developed to find all relevant candidate publications while limiting the number of unrelated ones. This method of publication discovery consists of four different steps:

##### 1. Publications linked at registration

ClinicalTrials.gov provides researchers with the opportunity to reference publications in the trial registration. These references may be related to the trial subject, or describe previous similar studies, or arise directly from the trial. While ClinicalTrials.gov contains metadata about the reference type (e.g. “results”), these metadata are unreliable. Thus, all referenced publications that could be found on PubMed were included. Conversely, related publications that could not be found in PubMed were disregarded.

##### 2. PubMed search

PubMed provides an API (“ESearch”) to access publication metadata and abstracts. An API is a standardized way of communication between software processes, often performed on the internet. The PubMed API was called using a search string which included the registration title, investigator names, publication date, and NCT-ID (Figure S1). The search string used is described in Figure S1. Since PubMed can return an arbitrary number of results in a single search, we limit the number of included publications to the 5 most relevant as decided by PubMed.

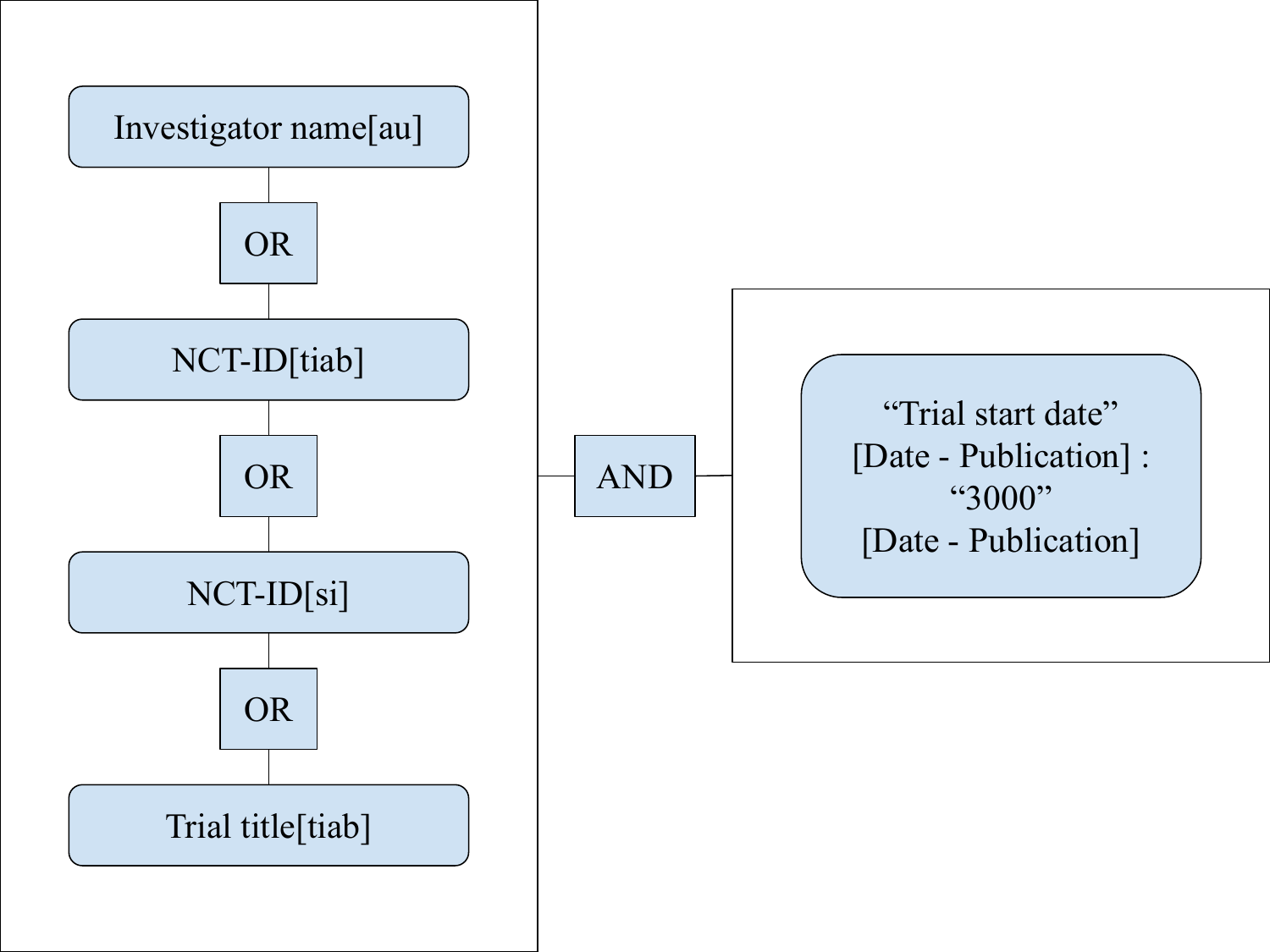

**Figure S1. Predefined PubMed search query.**

Boxes represent search term groupings (i.e., parentheses). Text terms are placeholders for their respective trial registration data. Terms in brackets are PubMed search field shorthand as follows; [tiab]: Title/Abstract; [si]: Secondary identifier; [au]: Author. “3000” represents the year 3000.

##### 3. LLM-enhanced PubMed search.

For this search strategy, the PubMed “ESearch” API was again called. However, instead of using a predefined search query, ChatGPT-5.1 (Version “gpt-5.1-2025-11-13”) was prompted to construct a query based on the trial registration. The two prompts included all registration data retrieved from ClinicalTrials.gov for the relevant trial. The generated search queries were then used to call the PubMed API, providing TrialScout with a ranked list of publications. The prompts used to create the search strings are available in Table S1.

**Table S1. LLM Prompts for query generation.**

Prompts used for ChatGPT-5.1 when constructing search queries. Grammatical errors have been preserved for reproducibility.

| Use-case | Prompt |
| --- | --- |
| To construct PubMed search queries for publication discovery #1: | You are a cunning pubmed expert, proficient in crafting search strings able to find publications relating to clinical trial registrations. You will be given information on a clinical trial from a clinical trial registry. You will craft a search string to find a given publication on pubmed. A good example of a search string would be:   ( NCTXXXXXXXX[si] OR  NCTXXXXXXXX[tiab] OR  "TITLE of the registration"[tiab] OR (keyword1[tiab] AND keyword2[tiab] AND keyword3[tiab])  )  AND  ("YYYY/MM/DD"[Date - Publication] : "3000"[Date - Publication])  AND  ( "Firstname Lastname"[Author] OR  "Another Author you found"[Affiliation] OR  "Name of institution/university"[Affiliation] )  Explanation for the dates: First date should be trial STUDY START. Second date should be "3000" to include all publications to the year 3000.  Affiliation field is good to use for both author and institution/university  Your input registration will be provided as JSON |
| To construct PubMed search queries for publication discovery #2: | You are an expert at finding publications from clinical trial registrations. You will be given a registration. Based on the registration you will produce short PubMed queries using this process:  1. Extract 2 pairs of keywords from the trial. Use as specific and short keywords as possible. Keywords are key aspects of the trial. For example: the disease (heart failure, osteosarcoma), drug names (imatinib, aspirin), trial acronyms (SAVE-IT, HF-REVEAL). 2. Find all people listed as investigators. If more than 3 are listed, select the 3 most important ones. 3. For each Investigator, construct 2 queries using the 2 pairs of keywords using the following format: Surname X[au] AND (<Keyword 1> OR <Keyword 2>) where X is the first letter of the persons first name. 4. Do not use quotation marks. 5. Do not use the NCT-ID as a keyword. 6. Finally - give one additional specific query that you think has the best chances of having the publication in top 5 search results. If no investigators are listed, provide 3 such queries instead. 7. Output your answer in the following JSON format:  { registration_nct_id: “NCTXXXXXXX” keywords: [“Keyword 1”, “Keyword 2”, “Keyword 3”, “Keyword 4”], investigators: [“<Investigator 1 first name and last name>”, “<Investigator 2 first and last name>”, “<Investigator 3 first and last name>”], search_strings: [“Surname1 X[au] AND (<Keyword 1> OR <Keyword 2>)”, “Surname X[au] AND (<Keyword 3> OR <Keyword 4>)”, “Surname2 X[au] AND (<Keyword 1> OR <Keyword 2>)”, “Surname2 X[au] AND (<Keyword 3> OR <Keyword 4>)”, “Surname3 X[au] AND (<Keyword 1> OR <Keyword 2>)”, “Surname3 X[au] AND (<Keyword 3> OR <Keyword 4>)”] extra_queries: [“<Your specific query(ies) from step 6>”]  } |

Abbreviations: JSON: JavaScript Object Notation (a structured text format)

*4. Google Scholar and PubMed Citation Match API*

Manual methods of discovery in previous studies in the field often incorporate Google searches to identify publications (1,2). Google provides many different search functions (e.g., Search, Images, Scholar, and more), however not all search results are useful. For example, using Google Search to search for publications using trial registration data could return links to the trial registration itself, or other links containing related or unrelated content. As opposed to this, Google Scholar always returns a list of publications, where the link title is always the publication title. While the links often lead to publisher’s websites, the corresponding publication could often also be found on PubMed. Unfortunately, Google doesn’t provide access to Google Scholar through an API, leaving us to rely on a third-party service (Serper.dev) to collect article titles from Google Scholar. The NCT-ID of the trial registration was used as the search term, as this was found to generate the most relevant matches. We hypothesize that Google Scholar indexes the full text of publications (the technical details are not disclosed by Google, to our knowledge). This indexing of the full text would explain the superior performance of this search strategy, as compared to NCT-ID match as previously described.

After retrieval of publication titles through Google Scholar, the PubMed Citation Matcher was used to identify the single best matching PubMed record, if any. The Citation Matcher’s “heuristic” retrieval method was used, searching with the publication title retrieved from Google Scholar. If a publication with the exact same title could be identified in our offline PubMed database, no request to the Citation Matcher API would be sent. If the Citation Matcher API did not provide any matching publication, a request would also be sent to the PubMed “ESearch” API with the publication title. Up to 10 publications arising from this search strategy were included, corresponding to the first page of Google Scholar search results.

##### Performance comparison of each search strategy

Table S2 details the number of candidate and result publications yielded by each search strategy. Notably, LLM-constructed PubMed search queries resulted in the highest number of both candidate and result publications. However, candidate publications found linked to from the trial registration were most likely to be classified as result publications, with a ratio of result to candidate publications of 72.2%.

**Table S2. Publications found per search strategy.**

Publications found by TrialScout during publication searches for 9,600 randomly sampled clinical trials. Percentages in parentheses represent the ratio of publications that were classified as belonging to that category. A single publication can be identified through multiple search strategies and thus belong to multiple categories. Percentages in the rightmost column represent the ratio of result publications per candidate publication for each search strategy.

| Strategy | Candidate publications | Result publications | Precision |
| --- | --- | --- | --- |
| Predefined PubMed search query | 21,154 (16.9%) | 3,247 (28.8%) | 15.3% |
| LLM-constructed PubMed search query | 93,419 (74.4%) | 8,698 (77.3%) | 9.3% |
| Linked to from trial registration | 7,430 (5.9%) | 5,366 (47.7%) | 72.2% |
| NCT-ID-search in Google Scholar | 22,647 (18%) | 6,733 (59.8%) | 29.7% |
| **All** | **125,528 (100%)** | **11,256 (100%)** | **9%** |

Abbreviations: LLM: Large Language Model; NCT-ID: National Clinical Trial Identifier.

#### Result detection

The process of publication discovery yielded a list of candidate publications, each of them potentially containing results of the trial. To determine whether a candidate publication did in fact contain results, we used the LLM ChatGPT-5.1 (3). We prompted the LLM with select fields from the trial registration as well as the title and abstract of a single publication. The data fields used to construct the prompt, and the prompt itself are available in Tables S3 and S4 respectively. To save costs and avoid rate-limits imposed by OpenAI, all prompts were sent in a single group, called a batch job, using an API. The temperature setting of the model was set to 1.0 and the reasoning effort was set to “medium”. After completion of the batch job, LLM predictions were merged with data of their respective publication and trial, effectively creating a list of result publications. If any result publications existed for a trial, it was marked as having detected results. Finally, data on examined trials were aggregated and exported in comma-separated values (csv) format for further analysis. The cost of running the requests averaged to 0.043 USD per analysed trial, varying greatly depending on the number of candidate publications found.

**Table S3. Data fields used to prompt LLM for result detection.**

| Trial data field | Explanation (When applicable) |
| --- | --- |
| **Trial registration data from ClinicalTrials.gov** |  |
| Trial title (brief) |  |
| Official trial title |  |
| Organization | The full name of the sponsor organization. |
| NCT-ID |  |
| Study type | Study type as listed in ClinicalTrials.gov, either interventional or observational. |
| Summary | A limited length summary of the trial. |
| Description | A detailed description of the trial. |
| **Publication data from PubMed** |  |
| Title |  |
| Abstract |  |
| Authors | All listed authors of a publication. |

Abbreviations: NCT-ID: National Clinical Trial number.

**Table S4. LLM Prompts for result detection.**

Prompts used for result detection using ChatGPT-5.1. Grammatical errors have been preserved for reproducibility.

| Use-case | Prompt |
| --- | --- |
| Determine whether a publication contains results of a given clinical trial registration | - You will act as a researcher.  - You will be given a registration for a clinical trial. The registration will contain various information fields describing the clinical trial, such as free text description, enrollment numbers, study design, and more.  - You will also receive a scientific article retrieved from PubMed. It contains the publication title, authors, and abstract. The publication may or may not relate to the clinical trial.  - Your task is to determine if the publication will contain the results of the previously given trial registration. You will output your best guess on whether this publication contains the results of said registration. - You will provide your judgement as a JSON object according to the schema provided to you  When attempting to match a registration to a publication, please consider the following: - Does the study design match? Consider randomization, groups, blinding, if the trial is multicenter, and so on. - Does the population match? Is the condition the same in both the registration and the publication? Is the same age group included? - Does the intervention match? Is the registration and publication actually examining the same intervention? - Is the enrollment numbers similar between registration and publication? It does not need to match exactly, but beware of discrepancies of high magnitude here. - Is the publication actually a meta-analysis? Meta-analysis may report results of many different studies but should not be deemed as having results for this registration. - Is the publication only a clinical trial protocol? Protocols do not contain any results and should therefore not be marked as having results. |

#### Future Developments

Further expansion of the tool’s scope is technically feasible as long as the sources contain text data in a known structured format. While TrialScout currently uses OpenAI’s API, the method is inherently agnostic to the underlying LLM and can be adapted to other LLMs that exist or will be available in the future. TrialScout could be further improved by implementing retrieval-augmented generation (4), chain-of-thought prompting (5), or by utilising a multi-LLM consensus workflow.

### Appendix 2 – Overview of currently available tools for automated results detection.

**Table S5. Existing automated tools for clinical trial result detection**

| Authors (Name) | Scope | Detection method | Evaluation |
| --- | --- | --- | --- |
| A. Powell-Smith, B. Goldacre, (TrialsTracker) (6) | Indexes all later phase interventional studies in ClinicalTrials.gov completed more than 24 months ago. Searches for results in ClinicalTrials.gov and explicit NCT-ID links in PubMed publications. | Algorithmic method based on regular expressions. | Derived sensitivity 69.8%, specificity 58.3% (vs. manual audit, n=2,562)¹ |
| N. Smalheiser, A. Holt (7) | Provides result searches for individual trials registered in ClinicalTrials.gov. Searches for matching PubMed publications. | Logistic regression model comparing similarity between trial-publication pairs based on metadata. | Recall 84.6%, precision 90.4%, AUC 0.95 (pair classification, n=13,042)² |
| T. Goodwin, M. Skinner, S. Harabagiu (8) | Links clinical trials in ClinicalTrials.gov to MEDLINE articles reporting their results. | Deep Highway Network with learning-to-rank features from trial and article metadata. | MAP 0.31, MRR 0.34 (closed); MAP 0.82, MRR 0.87 (open)³ |

1: Derived from comparison with manual audit by Chen et al. (n=2,562 overlapping trials). Not reported by the original authors.

2: Metrics are for trial-publication pair classification on a constructed balanced hold-out test set (n=13,042).

3: Information retrieval ranking metrics (Mean Average Precision and Mean Reciprocal Rank). “Closed” uses only registry metadata; “open” also uses linked publications.
