## Appendix 3 for "TrialScout links published results to trial registrations using a large language model"

### Appendix 3. STROBE checklist

STROBE Statement, checklist of items that should be included in reports of cross-sectional studies. Source: von Elm E, Altman DG, Egger M, Pocock SJ, Gøtzsche PC, Vandenbroucke JP, et al. The Strengthening the Reporting of Observational Studies in Epidemiology (STROBE) statement: guidelines for reporting observational studies. J Clin Epidemiol. 2008 Apr;61(4):344–9. doi:10.1016/j.jclinepi.2007.11.008 PubMed PMID: 18313558.

|  | **Item No** | **Recommendation** | **Reported in** |
| --- | --- | --- | --- |
| Title and abstract | 1 | (a) Indicate the study’s design with a commonly used term in the title or the abstract | Abstract |
|  |  | (b) Provide in the abstract an informative and balanced summary of what was done and what was found | Abstract |
| **Introduction** |  |  |  |
| Background/rationale | 2 | Explain the scientific background and rationale for the investigation being reported | Section 1 |
| Objectives | 3 | State specific objectives, including any prespecified hypotheses | Section 1, paragraph 4; Section 2.4, paragraph 2 |
| **Methods** |  |  |  |
| Study design | 4 | Present key elements of study design early in the paper | Section 2.4, paragraph 1 |
| Setting | 5 | Describe the setting, locations, and relevant dates, including periods of recruitment, exposure, follow-up, and data collection | Section 2.4, paragraphs 1 and 2; Figure 2 |
| Participants | 6 | (a) Give the eligibility criteria, and the sources and methods of selection of participants | Section 2.4, paragraph 1; Figure 2 |
| Variables | 7 | Clearly define all outcomes, exposures, predictors, potential confounders, and effect modifiers. Give diagnostic criteria, if applicable | Section 2.1; Section 2.4, paragraph 2 |
| Data sources/ measurement | 8 | For each variable of interest, give sources of data and details of methods of assessment (measurement). Describe comparability of assessment methods if there is more than one group | Sections 2.2 and 2.3; Section 2.4, paragraphs 1 and 2; Section 3.1 |
| Bias | 9 | Describe any efforts to address potential sources of bias | Section 2.3, paragraphs 2 and 4; Section 2.4, paragraph 2; Section 4.1 |
| Study size | 10 | Explain how the study size was arrived at | Section 2.4, paragraph 1 |
| Quantitative variables | 11 | Explain how quantitative variables were handled in the analyses. If applicable, describe which groupings were chosen and why | Section 2.4, paragraph 2; Table 2; Figure 4 |
| Statistical methods | 12 | (a) Describe all statistical methods, including those used to control for confounding | Section 2.4, paragraph 2 |
|  |  | (b) Describe any methods used to examine subgroups and interactions | Section 2.4, paragraph 2. No interactions were tested |
|  |  | (c) Explain how missing data were addressed | Table 2 gives the missing counts. Handling in the analyses is not stated |
|  |  | (d) If applicable, describe analytical methods taking account of sampling strategy | Section 2.4, paragraph 1. Simple random sample, no weighting applied |
|  |  | (e) Describe any sensitivity analyses | None performed |
| **Results** |  |  |  |
| Participants | 13 | (a) Report numbers of individuals at each stage of study | Section 2.4, paragraph 1; Figure 2 |
|  |  | (b) Give reasons for non-participation at each stage | Figure 2 |
|  |  | (c) Consider use of a flow diagram | Figure 2 |
| Descriptive data | 14 | (a) Give characteristics of study participants and information on exposures and potential confounders | Section 3.2; Table 2 |
|  |  | (b) Indicate number of participants with missing data for each variable of interest | Table 2 |
| Outcome data | 15 | Report numbers of outcome events or summary measures | Section 3.3, paragraph 1; Figure 3; Table 3 |
| Main results | 16 | (a) Give unadjusted estimates and, if applicable, confounder-adjusted estimates and their precision. Make clear which confounders were adjusted for and why they were included | Section 3.3, paragraph 2; Table 3; Figure 4. All estimates are unadjusted |
|  |  | (b) Report category boundaries when continuous variables were categorized | Table 2; Figures 4 and 5 |
|  |  | (c) If relevant, consider translating estimates of relative risk into absolute risk for a meaningful time period | Section 3.3, paragraph 2; Table 3 |
| Other analyses | 17 | Report other analyses done, for example analyses of subgroups and interactions, and sensitivity analyses | Section 3.3, paragraph 2; Table 3 |
| **Discussion** |  |  |  |
| Key results | 18 | Summarise key results with reference to study objectives | Section 4, paragraph 1; Section 5 |
| Limitations | 19 | Discuss limitations of the study, taking into account sources of potential bias or imprecision. Discuss both direction and magnitude of any potential bias | Section 4.1; Section 4.2, paragraph 3 |
| Interpretation | 20 | Give a cautious overall interpretation of results considering objectives, limitations, multiplicity of analyses, results from similar studies, and other relevant evidence | Sections 4 to 4.3; Section 5 |
| Generalisability | 21 | Discuss the generalisability (external validity) of the study results | Section 4.1, paragraph 1; Section 4.2, paragraph 2 |
| **Other information** |  |  |  |
| Funding | 22 | Give the source of funding and the role of the funders for the present study and, if applicable, for the original study on which the present article is based | Declarations, Funding |
