## Appendix 4 for "TrialScout links published results to trial registrations using a large language model"

### Appendix 4. Subgroup analyses on all three outcome measures

Subgroup analyses of the 9,600 randomly sampled trials, repeated on each of the three outcome measures defined in Section 2.1. Published results are those found in a peer-reviewed publication. Summary results are those posted to ClinicalTrials.gov. Reported results are either of the two. Table 3 in the main text reports significance on published results.

**Table A4.1. Results reporting by trial characteristic, n/N (%).**

| **Subgroup** | **Reported results** | **p** | **Published results** | **p** | **Summary results** | **p** |
| --- | --- | --- | --- | --- | --- | --- |
| **Study status** |  | <0.001 |  | <0.001 |  | <0.001 |
| Completed | 6407/8561 (74.8%) |  | 5816/8561 (67.9%) |  | 2307/8561 (26.9%) |  |
| Terminated | 591/1039 (56.9%) |  | 294/1039 (28.3%) |  | 436/1039 (42.0%) |  |
| **Phase** |  | <0.001 |  | <0.001 |  | <0.001 |
| Early Phase 1 | 64/104 (61.5%) |  | 59/104 (56.7%) |  | 13/104 (12.5%) |  |
| Phase 1 | 784/1301 (60.3%) |  | 691/1301 (53.1%) |  | 223/1301 (17.1%) |  |
| Phase 1/Phase 2 | 264/331 (79.8%) |  | 204/331 (61.6%) |  | 161/331 (48.6%) |  |
| Phase 2 | 1176/1505 (78.1%) |  | 933/1505 (62.0%) |  | 707/1505 (47.0%) |  |
| Phase 2/Phase 3 | 119/167 (71.3%) |  | 108/167 (64.7%) |  | 47/167 (28.1%) |  |
| Phase 3 | 921/1095 (84.1%) |  | 812/1095 (74.2%) |  | 527/1095 (48.1%) |  |
| Phase 4 | 714/915 (78.0%) |  | 598/915 (65.4%) |  | 315/915 (34.4%) |  |
| Missing/Not applicable | 2956/4182 (70.7%) |  | 2705/4182 (64.7%) |  | 750/4182 (17.9%) |  |
| **Lead Sponsor Type** |  | <0.001 |  | <0.001 |  | <0.001 |
| Other | 4504/6116 (73.6%) |  | 4129/6116 (67.5%) |  | 1269/6116 (20.7%) |  |
| Industry | 2199/3120 (70.5%) |  | 1725/3120 (55.3%) |  | 1320/3120 (42.3%) |  |
| U.S. Federal Agency/NIH | 295/364 (81.0%) |  | 256/364 (70.3%) |  | 154/364 (42.3%) |  |
| **Participant Sex** |  | 0.097 |  | 0.641 |  | <0.001 |
| All | 5951/8119 (73.3%) |  | 5189/8119 (63.9%) |  | 2407/8119 (29.6%) |  |
| Female only | 658/914 (72.0%) |  | 567/914 (62.0%) |  | 230/914 (25.2%) |  |
| Male only | 385/561 (68.6%) |  | 350/561 (62.4%) |  | 105/561 (18.7%) |  |
| Missing | 4/6 (66.7%) |  | 4/6 (66.7%) |  | 1/6 (16.7%) |  |

p values are from Pearson’s chi-squared tests on the full contingency table for each characteristic. Percentages are of the trials in that row. Published and summary results are not mutually exclusive.

**Table A4.2. Prespecified comparisons, difference in percentage points.**

| **Comparison** | **Outcome** | **Group of interest** | **Comparator** | **Difference (pp)** | **95% CI** | **p** |
| --- | --- | --- | --- | --- | --- | --- |
| Industry vs non-industry | Reported results | 2199/3120 (70.5%) | 4799/6480 (74.1%) | -3.6 | -5.5 to -1.6 | <0.001 |
|  | Published results | 1725/3120 (55.3%) | 4385/6480 (67.7%) | -12.4 | -14.5 to -10.3 | <0.001 |
|  | Summary results | 1320/3120 (42.3%) | 1423/6480 (22.0%) | 20.3 | 18.3 to 22.4 | <0.001 |
| Male-only vs all other trials | Reported results | 385/561 (68.6%) | 6609/9033 (73.2%) | -4.5 | -8.6 to -0.5 | 0.022 |
|  | Published results | 350/561 (62.4%) | 5756/9033 (63.7%) | -1.3 | -5.6 to 2.9 | 0.554 |
|  | Summary results | 105/561 (18.7%) | 2637/9033 (29.2%) | -10.5 | -13.9 to -7.0 | <0.001 |
| Early phase vs later phase | Reported results | 848/1405 (60.4%) | 3194/4013 (79.6%) | -19.2 | -22.1 to -16.3 | <0.001 |
|  | Published results | 750/1405 (53.4%) | 2655/4013 (66.2%) | -12.8 | -15.8 to -9.7 | <0.001 |
|  | Summary results | 236/1405 (16.8%) | 1757/4013 (43.8%) | -27.0 | -29.5 to -24.5 | <0.001 |

Differences are the group of interest minus the comparator, computed on unrounded proportions. Confidence intervals are from a test of two proportions, p values from Pearson’s chi-squared test. Phase comparisons exclude trials with no phase recorded, and sex comparisons exclude trials with no sex recorded. In each comparison the group of interest is named first. Early phase comprises early phase 1 and phase 1.
